# Efficacy and Safety of Bempedoic Acid in Patients With and Without Metabolic Syndrome: Pooled Analysis of Data From Four Phase 3 Clinical Trials

**DOI:** 10.1101/2023.03.03.23286700

**Authors:** Michael D. Shapiro, Pam R. Taub, Michael J. Louie, Lei Lei, Christie M. Ballantyne

## Abstract

**Background and aims:** Bempedoic acid significantly lowers low-density lipoprotein cholesterol (LDL-C) in patients with hypercholesterolemia but its effects in patients with metabolic syndrome (MetS) have not been well characterized. We sought to determine the efficacy and safety of bempedoic acid in patients with hypercholesterolemia by baseline MetS status.

**Methods:** This study used pooled data from four phase 3 studies. Using modified International Atherosclerosis Society guidelines, patients were grouped into two pools: those with and those without MetS. Patients with diabetes were excluded. Endpoints assessed change from baseline to week 12 in lipid and glycemic parameters and high-sensitivity C-reactive protein (hsCRP), and safety.

**Results:** The study included 936 patients with MetS (bempedoic acid, 648; placebo, 288) and 1573 without MetS (bempedoic acid, 1037; placebo, 536). Significant placebo-corrected reductions in LDL-C were observed with bempedoic acid (*p*<0.0001), with a greater decrease in patients with *vs*. without MetS (−22.3% *vs*. −18.4%; interaction *p*=0.0472). Compared with placebo, bempedoic acid significantly (*p*<0.0001) lowered total cholesterol, non–high-density lipoprotein cholesterol, apolipoprotein B, and hsCRP, with a similar magnitude of benefit observed between MetS categories. Significant reductions in triglycerides (−4.4%; *p*=0.02) were only observed in patients without MetS; only patients with MetS experienced decreases in glycated hemoglobin (−0.07%; *p*<0.0001) and fasting plasma glucose (−2.4 mg/dL; *p*=0.002). Safety was comparable between MetS categories and treatment groups.

**Conclusions:** These data suggest that bempedoic acid is a suitable therapy for patients with MetS who require additional lipid lowering.

**Highlights:**

- Bempedoic acid lowered LDL-C *vs*. placebo in patients with and without MetS.
- Placebo-corrected LDL-C reductions were greater in patients with *vs*. without MetS.
- Bempedoic acid lowered TC, non–HDL-C, ApoB, and hsCRP regardless of MetS category.
- Reductions in HbA1c and FPG *vs*. placebo were only observed in patients with MetS.
- Bempedoic acid safety was comparable between MetS categories.

## Introduction

Metabolic syndrome (MetS) is defined as the co-occurrence of a group of physiologic and metabolic abnormalities that are risk factors for atherosclerotic cardiovascular disease (ASCVD) and new-onset diabetes [1, 2]. MetS is associated with a 2-fold increase in risk of developing ASCVD and a 5-fold increase in risk of new-onset diabetes [1]. Risk factors for MetS include hyperglycemia, dyslipidemia, hypertension, and obesity. Hyperglycemia contributes to the development of cardiotoxicity and ASCVD by inducing endothelial and myocardial cell dysfunction, inflammation, and proatherogenic cellular and molecular changes [3].

Glycemic control has been shown to be improved by certain lipid-lowering therapies (LLTs), including bile acid sequestrants [4] and fibrates [5]; however, not all LLTs are associated with such improvements. Statins, which are recommended as first-line treatment of patients with ASCVD to reduce cardiovascular risk [6], are associated with elevated glycemic parameters and increased risk of developing diabetes [7-11]; similar associations have been observed for niacin [12, 13]. In contrast, the proprotein convertase subtilisin/kexin type 9 (PCSK9) inhibitors alirocumab, evolocumab, and inclisiran have little effect on glycemic parameters [14-16] or the incidence of new-onset diabetes [15, 17, 18].

Bempedoic acid is an oral, first-in-class ATP-citrate lyase inhibitor that has been shown to significantly lower low-density lipoprotein cholesterol (LDL-C) compared with placebo in four phase 3 clinical trials [19-22]. In an analysis of pooled data from these four trials, treatment with bempedoic acid was associated with a significant 18–25% decrease in levels of LDL-C compared with placebo in patients with hypercholesterolemia receiving maximally tolerated statins [23]. Bempedoic acid was also associated with greater decreases in levels of total cholesterol, non–high-density lipoprotein cholesterol (non–HDL-C), apolipoprotein B (ApoB), and high-sensitivity C-reactive protein (hsCRP) compared with placebo [23].

A separate analysis using pooled data from the same four phase 3 trials assessed the efficacy and safety of bempedoic acid in patients with diabetes, prediabetes, and normoglycemia at baseline [24]. Results from this study showed that compared with placebo, bempedoic acid significantly lowered LDL-C across all three glycemic status groups and glycated hemoglobin (HbA1c) in patients with diabetes or prediabetes.

The effect of bempedoic acid as a LLT in patients with MetS has not yet been fully established. In this study, we investigated the efficacy and safety of bempedoic acid, and its effect on glycemic parameters, by baseline metabolic status.

## Patients and methods

### Study Design

This study is a post-hoc analysis using pooled data from four phase 3 randomized, double-blind, placebo-controlled, parallel-group, multicenter studies of bempedoic acid. The details of these studies, CLEAR Harmony (NCT02666664) [22], CLEAR Wisdom (NCT02991118) [20], CLEAR Serenity (NCT02988115) [21], and CLEAR Tranquility (NCT03001076) [19], have been previously reported.

All four studies enrolled adult patients with hypercholesterolemia receiving stable LLT who required additional lowering of LDL-C. Patients were randomized 2:1 to receive bempedoic acid 180 mg or placebo once daily. CLEAR Harmony and CLEAR Wisdom were 52-week studies that enrolled patients with established

ASCVD and/or heterozygous familial hypercholesterolemia who were receiving stable maximally tolerated statins alone or in combination with other LLT [20, 22]. The 24-week CLEAR Serenity and 12-week CLEAR Tranquility studies enrolled patients with a history of statin intolerance, who were receiving no greater than very low–dose or low-dose statin, respectively [19, 21]. All studies were conducted in accordance with the Declaration of Helsinki and Good Clinical Practice guidelines, and their respective protocols received approval from local independent ethics committees at each study site. All study participants provided written informed consent.

For the current analysis, patients with diabetes (those with a history of diabetes, baseline HbA1c ≥6.5%, or fasting plasma glucose [FPG] ≥126 mg/dL at both screening and randomization) were excluded. The remaining patients were grouped by baseline metabolic status into two categories: patients with MetS and those without MetS. MetS was defined using modified International Atherosclerosis Society guidelines [25], which require the presence of three or more of the following criteria: increased waist circumference, triglycerides ≥150 mg/dL, HDL-C <40 mg/dL for men or <50 mg/dL for women, history of hypertension or blood pressure ≥130/85 mmHg, and FPG ≥100 mg/dL. Body mass index (BMI) was used as a proxy for waist circumference, with BMI >25 kg/m^2^ and >30 kg/m^2^ for Asian and non-Asian patients, respectively, used to define increased waist circumference.

### Endpoints and Assessments

Prespecified endpoints in all four studies were percent change in LDL-C, total cholesterol, non–HDL-C, ApoB, hsCRP, and triglycerides from baseline to week 12, and absolute changes in FPG and HbA1c from baseline to week 12. LDL-C values were calculated using the Friedewald equation [26] or measured directly using the Multigent Direct LDL assay (Architect system, Abbott) when triglyceride levels were >400 mg/dL or LDL-C levels were <50 mg/dL. Quantification of lipids and biomarkers was performed at a central laboratory (Q^2^ Solutions).

Evaluations of the safety of bempedoic acid included investigator-reported treatment-emergent adverse events (TEAEs) and serious TEAEs.

### Statistical Analysis

The efficacy analysis population included all randomized patients, and the safety analysis population included all patients who received at least one dose of study treatment.

Least squares (LS) means, 95% confidence intervals (CIs), and *p* values for treatment differences within each MetS status category were based on an analysis of covariance (ANCOVA) model with percent change or change from baseline as the dependent variable, study and treatment as fixed factors, and baseline as a covariate. Comparisons of hsCRP were based on a nonparametric method. The interaction *p* value was obtained from the interaction term via an ANCOVA model with percent change or change from baseline as the dependent variable (log-transformed for hsCRP), study, treatment, MetS status, and the interaction term between treatment and MetS status as fixed factors, and baseline as a covariate. No multiplicity adjustment was performed. Adverse events were coded per the Medical Dictionary for Regulatory Activities (MedDRA) version 20.1 and summarized as patient incidences.

## Results

### Patients

Of the 3623 patients randomized to bempedoic acid or placebo across all four phase 3 studies, 1114 (30.7%) had diabetes at baseline and were excluded from these analyses. Among the remaining 2509 patients, there were 936 (37.3%) patients with MetS (648 [25.8%] and 288 [11.5%] randomized to bempedoic acid and placebo, respectively) and 1573 (62.7%) without MetS (1037 [41.3%] and 536 [21.4%] randomized to bempedoic acid and placebo, respectively; **Table 1**).

**Table 1.**
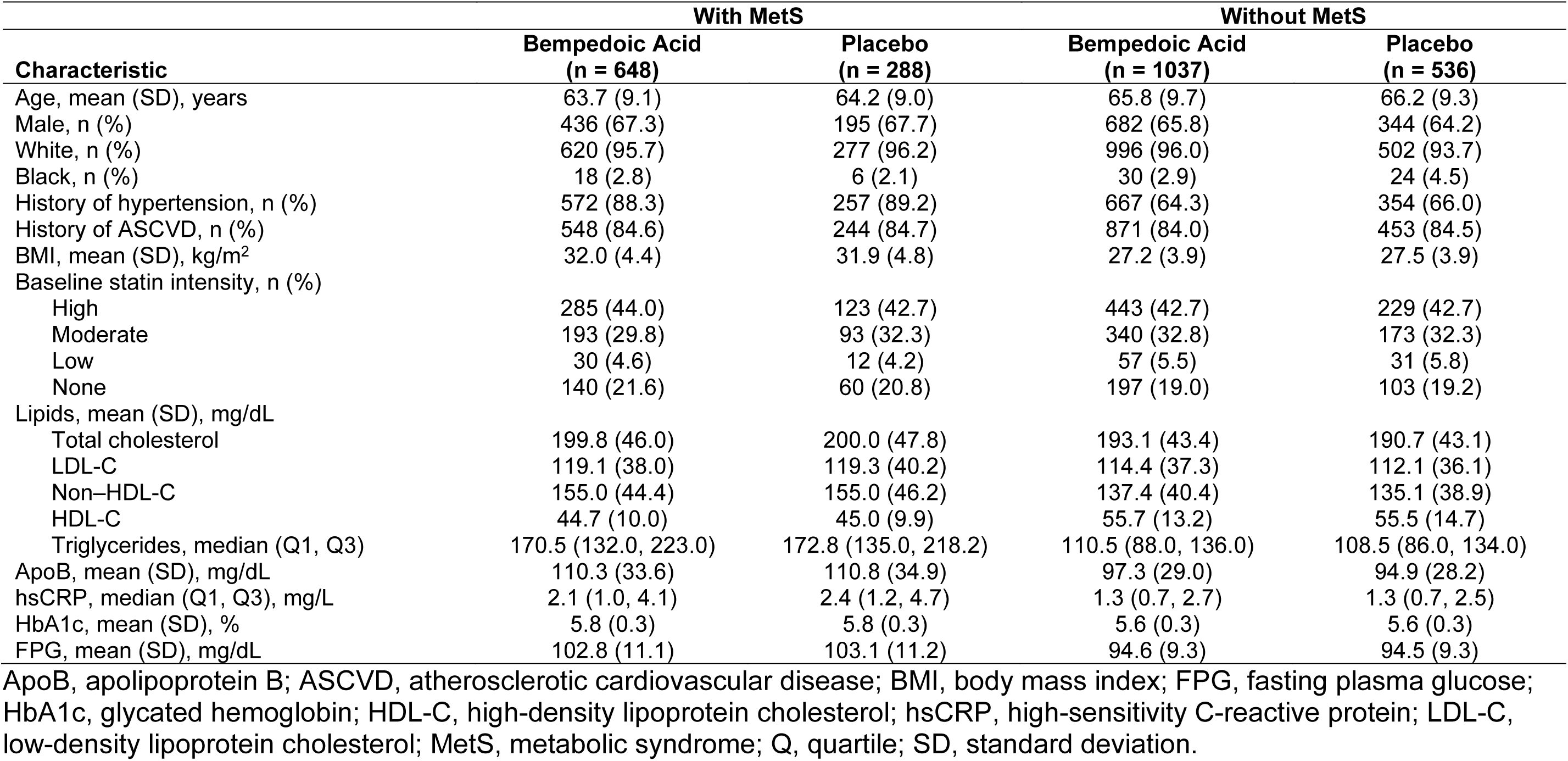
Baseline Demographics and Clinical Characteristics.

| Characteristic | With MetS |  | Without MetS |  |
| --- | --- | --- | --- | --- |
|  | Bempedoic Acid<br>(n = 648) | Placebo<br>(n = 288) | Bempedoic Acid<br>(n = 1037) | Placebo<br>(n = 536) |
| Age, mean (SD), years | 63.7 (9.1) | 64.2 (9.0) | 65.8 (9.7) | 66.2 (9.3) |
| Male, n (%) | 436 (67.3) | 195 (67.7) | 682 (65.8) | 344 (64.2) |
| White, n (%) | 620 (95.7) | 277 (96.2) | 996 (96.0) | 502 (93.7) |
| Black, n (%) | 18 (2.8) | 6 (2.1) | 30 (2.9) | 24 (4.5) |
| History of hypertension, n (%) | 572 (88.3) | 257 (89.2) | 667 (64.3) | 354 (66.0) |
| History of ASCVD, n (%) | 548 (84.6) | 244 (84.7) | 871 (84.0) | 453 (84.5) |
| BMI, mean (SD), kg/m<sup>2</sup> | 32.0 (4.4) | 31.9 (4.8) | 27.2 (3.9) | 27.5 (3.9) |
| Baseline statin intensity, n (%) |  |  |  |  |
| High | 285 (44.0) | 123 (42.7) | 443 (42.7) | 229 (42.7) |
| Moderate | 193 (29.8) | 93 (32.3) | 340 (32.8) | 173 (32.3) |
| Low | 30 (4.6) | 12 (4.2) | 57 (5.5) | 31 (5.8) |
| None | 140 (21.6) | 60 (20.8) | 197 (19.0) | 103 (19.2) |
| Lipids, mean (SD), mg/dL |  |  |  |  |
| Total cholesterol | 199.8 (46.0) | 200.0 (47.8) | 193.1 (43.4) | 190.7 (43.1) |
| LDL-C | 119.1 (38.0) | 119.3 (40.2) | 114.4 (37.3) | 112.1 (36.1) |
| Non-HDL-C | 155.0 (44.4) | 155.0 (46.2) | 137.4 (40.4) | 135.1 (38.9) |
| HDL-C | 44.7 (10.0) | 45.0 (9.9) | 55.7 (13.2) | 55.5 (14.7) |
| Triglycerides, median (Q1, Q3) | 170.5 (132.0, 223.0) | 172.8 (135.0, 218.2) | 110.5 (88.0, 136.0) | 108.5 (86.0, 134.0) |
| ApoB, mean (SD), mg/dL | 110.3 (33.6) | 110.8 (34.9) | 97.3 (29.0) | 94.9 (28.2) |
| hsCRP, median (Q1, Q3), mg/L | 2.1 (1.0, 4.1) | 2.4 (1.2, 4.7) | 1.3 (0.7, 2.7) | 1.3 (0.7, 2.5) |
| HbA1c, mean (SD), % | 5.8 (0.3) | 5.8 (0.3) | 5.6 (0.3) | 5.6 (0.3) |
| FPG, mean (SD), mg/dL | 102.8 (11.1) | 103.1 (11.2) | 94.6 (9.3) | 94.5 (9.3) |

ApoB, apolipoprotein B; ASCVD, atherosclerotic cardiovascular disease; BMI, body mass index; FPG, fasting plasma glucose; HbA1c, glycated hemoglobin; HDL-C, high-density lipoprotein cholesterol; hsCRP, high-sensitivity C-reactive protein; LDL-C, low-density lipoprotein cholesterol; MetS, metabolic syndrome; Q, quartile; SD, standard deviation.

Baseline demographics and laboratory parameters were generally comparable between the bempedoic acid and placebo treatment groups, both within MetS status categories and, with the exception of parameters included in MetS criteria, between MetS status categories (**Table 1**). Across all four groups, mean age was 63.7–66.2 years, approximately two-thirds of patients were male, and most were White (≥93.7%). Approximately 85% of patients across all groups had a history of ASCVD. Differences observed between patients with and without MetS were expected.

### Change in Lipid Parameters, hsCRP, and Glycemic Parameters

Significant reductions in LDL-C from baseline to week 12 were observed among patients who received bempedoic acid compared with those who received placebo, regardless of MetS status (*p*<0.0001; **Figure 1**). The placebo-corrected mean percent reduction in LDL-C was significantly more pronounced in patients with MetS than in those without MetS (−22.3% *vs*. −18.4%; interaction *p*=0.0472).

**Figure 1.**
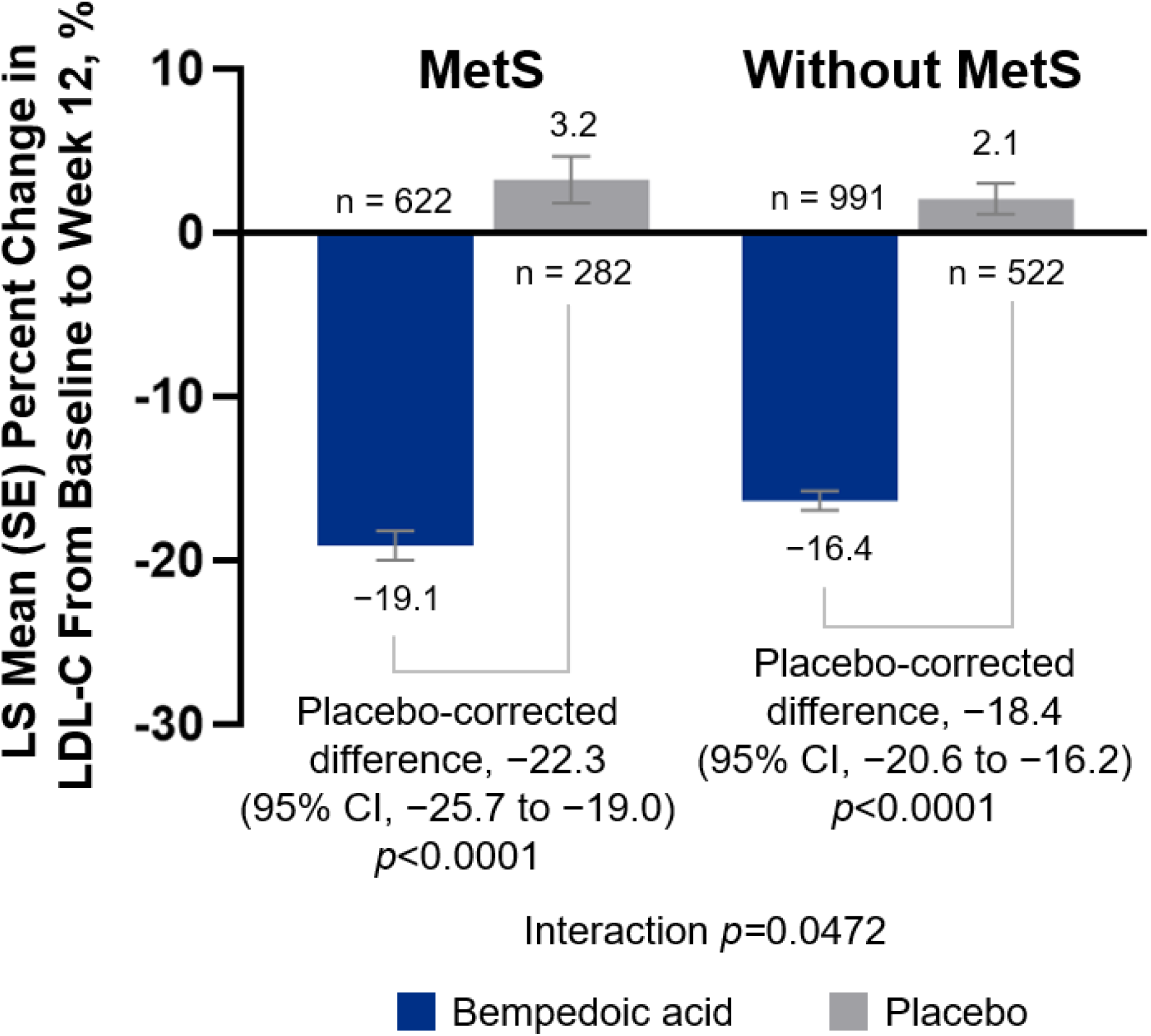
Change in LDL-C From Baseline to Week 12 in Patients With and Without MetS. CI, confidence interval; LDL-C, low-density lipoprotein cholesterol; LS, least squares; MetS, metabolic syndrome; SE, standard error. Numbers on the x axis represent the total number of patients within each group with available LDL-C data at week 12.

Within both MetS categories, significant reductions in total cholesterol, non–HDL-C, and ApoB from baseline to week 12 were observed in patients treated with bempedoic acid compared with those who received placebo (*p*<0.0001 for all comparisons; **Figure 2A–C**); the magnitude of benefit with bempedoic acid was similar between patients with and without MetS. Triglyceride levels increased from baseline to week 12 in patients treated with bempedoic acid, regardless of MetS status. However, in patients with MetS, triglyceride levels at week 12 were higher with bempedoic acid than placebo by 13.1% (*p*<0.0001), whereas in those without MetS, week-12 triglyceride levels were lower with bempedoic acid than placebo by 4.4% (*p*=0.02); the interaction was significant (*p*<0.0001; **Figure 2D**). Significant placebo-corrected reductions in hsCRP from baseline to week 12 with bempedoic acid were observed regardless of metabolic status (*p*<0.0001), with a similar magnitude of benefit in both MetS categories (**Figure 2E**). In patients with MetS, significant reductions from baseline to week 12 in HbA1c (*p*<0.0001; **Figure 2F**) and FPG (*p*=0.002; **Figure 2G**) were observed in patients treated with bempedoic acid compared with those who received placebo; no such differences were observed in patients without MetS (HbA1c, interaction *p*=0.0003; FPG, interaction *p*=0.002). Within each MetS status category, the change in mean body weight from baseline was ≤1.1% at every assessed time point over 52 weeks, regardless of treatment (**Figure 3**).

**Figure 2.**
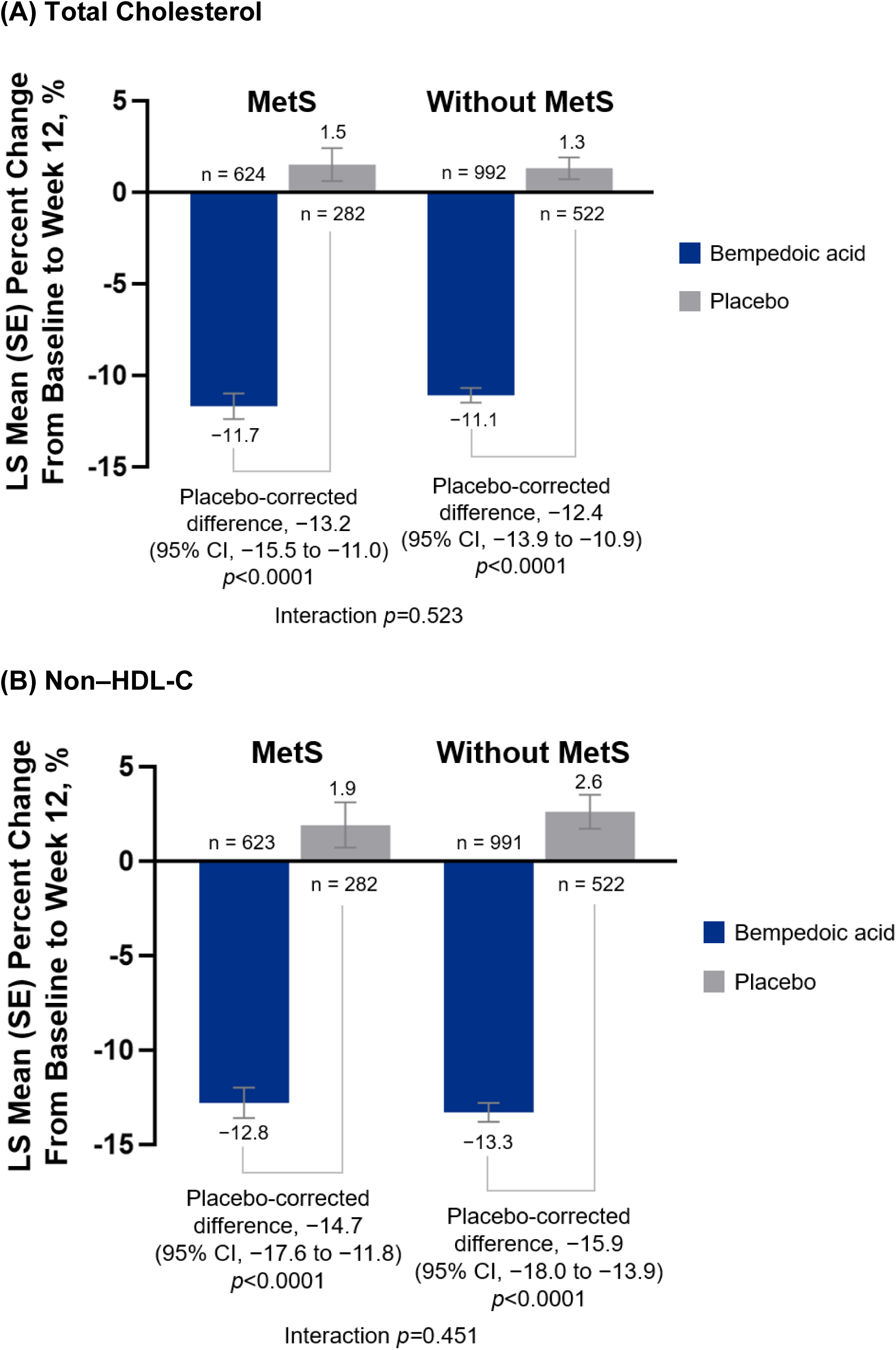
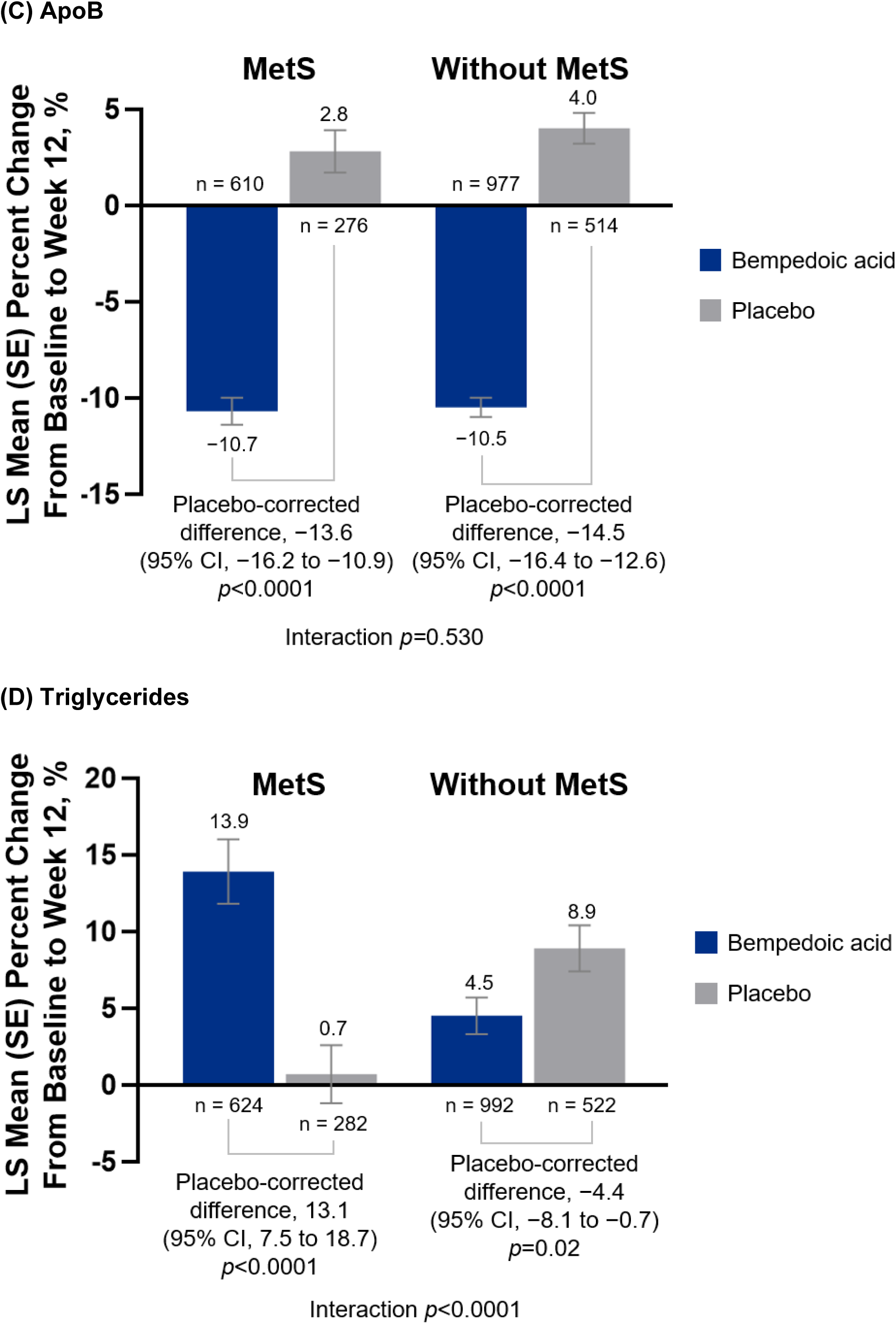
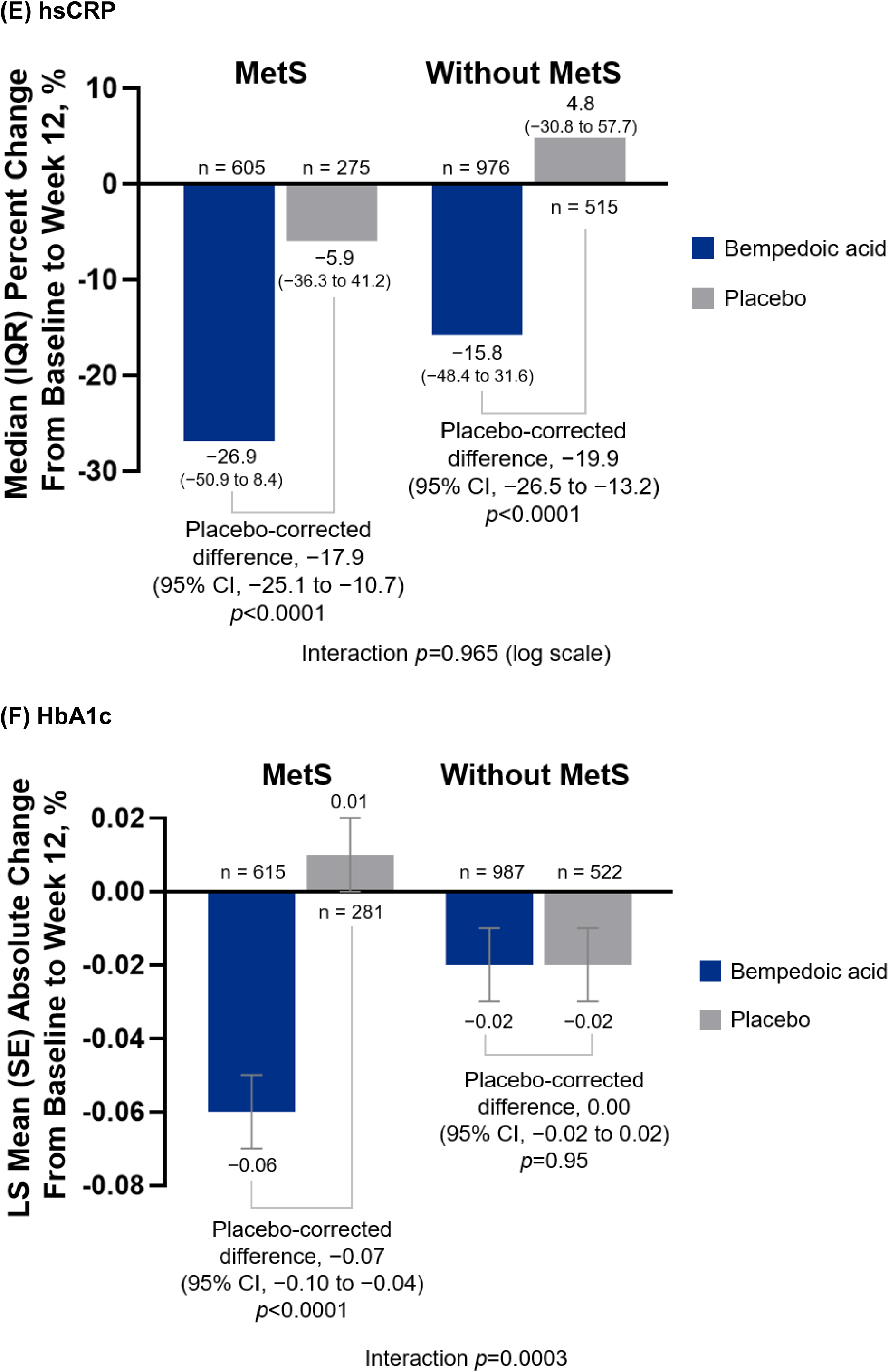
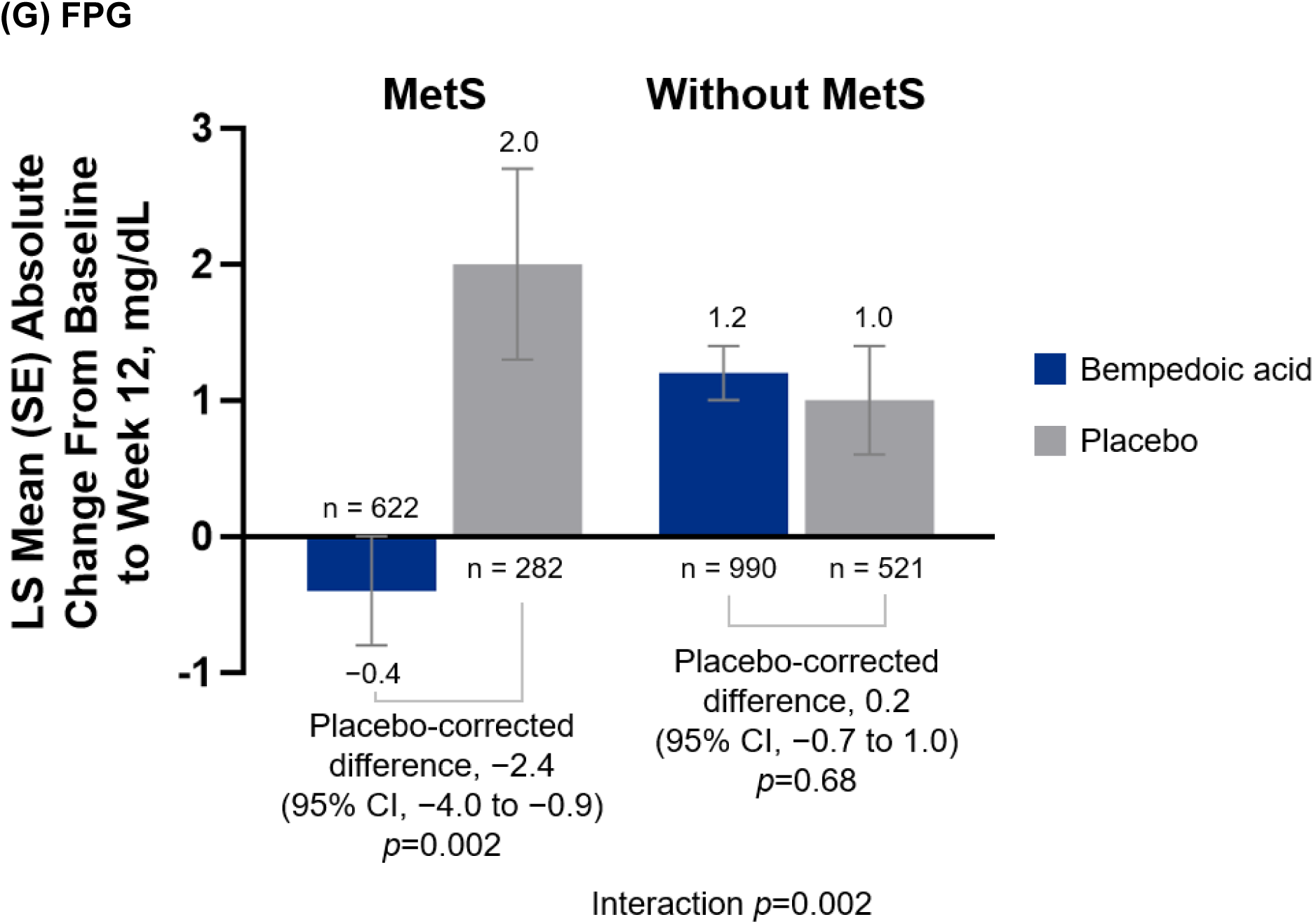
Change in Lipids, hsCRP, and Glycemic Parameters From Baseline to Week 12. ApoB, apolipoprotein B; CI, confidence interval; FPG, fasting plasma glucose; HbA1c, glycated hemoglobin; hsCRP, high-sensitivity C-reactive protein; IQR, interquartile range; LS, least squares; MetS, metabolic syndrome; non–HDL-C; non–high-density lipoprotein cholesterol; SE, standard error. Numbers on the x axis represent the total number of patients within each group with available data at week 12 for each parameter.

**Figure 3.**
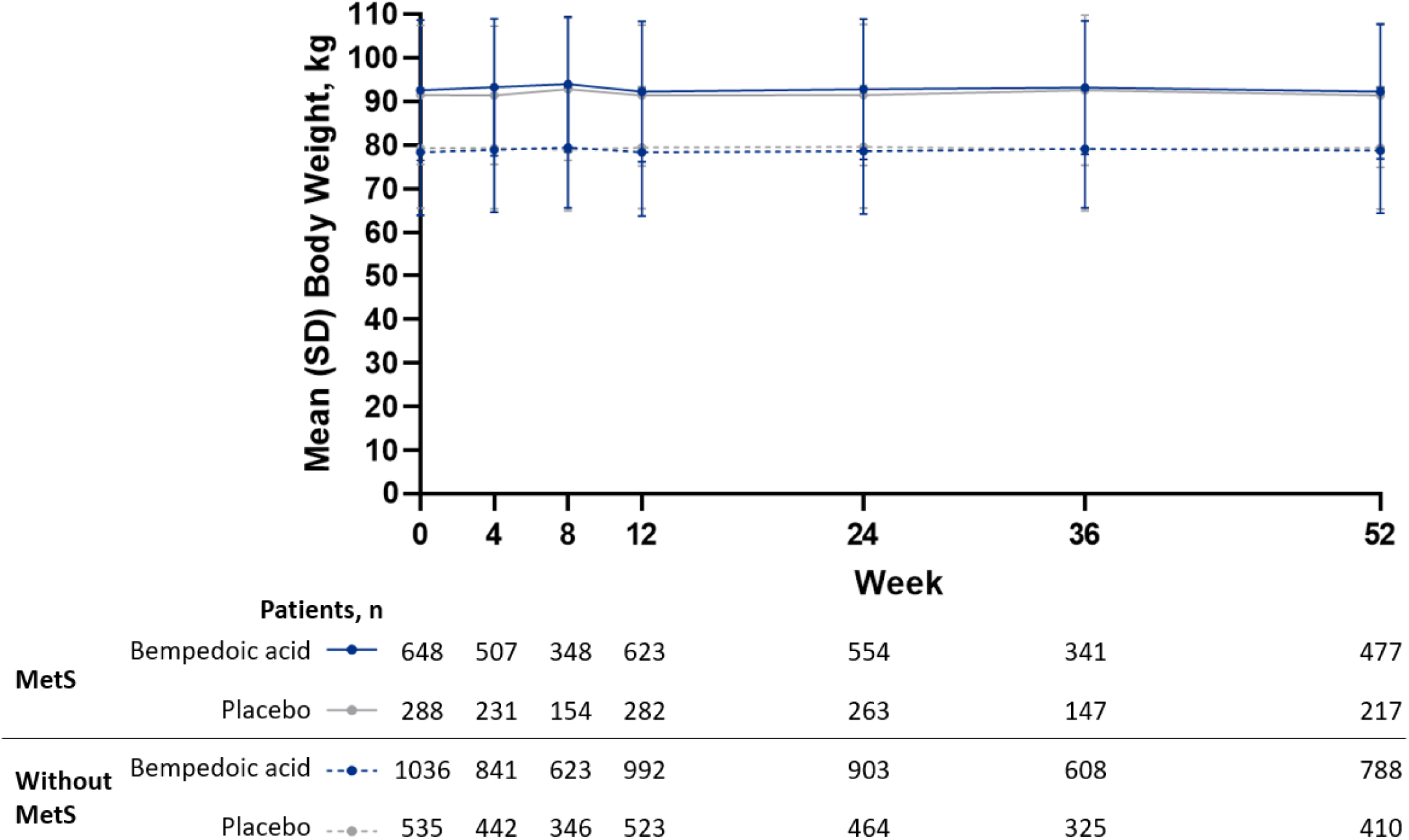
Body Weight Over Time. MetS, metabolic syndrome; SD, standard deviation.

### Safety

The safety profile of bempedoic acid was generally comparable between patients with and without MetS (**Table 2**). Among patients with MetS, TEAEs were reported in 459 (70.8%) patients treated with bempedoic acid and 194 (67.4%) patients who received placebo. Among patients without MetS, TEAEs were reported in 774 (74.7%) and 389 (72.7%) patients who received bempedoic acid and placebo, respectively. Serious TEAEs were reported in 81 (12.5%) patients with MetS treated with bempedoic acid and 36 (12.5%) patients with MetS who received placebo.

**Table 2.**
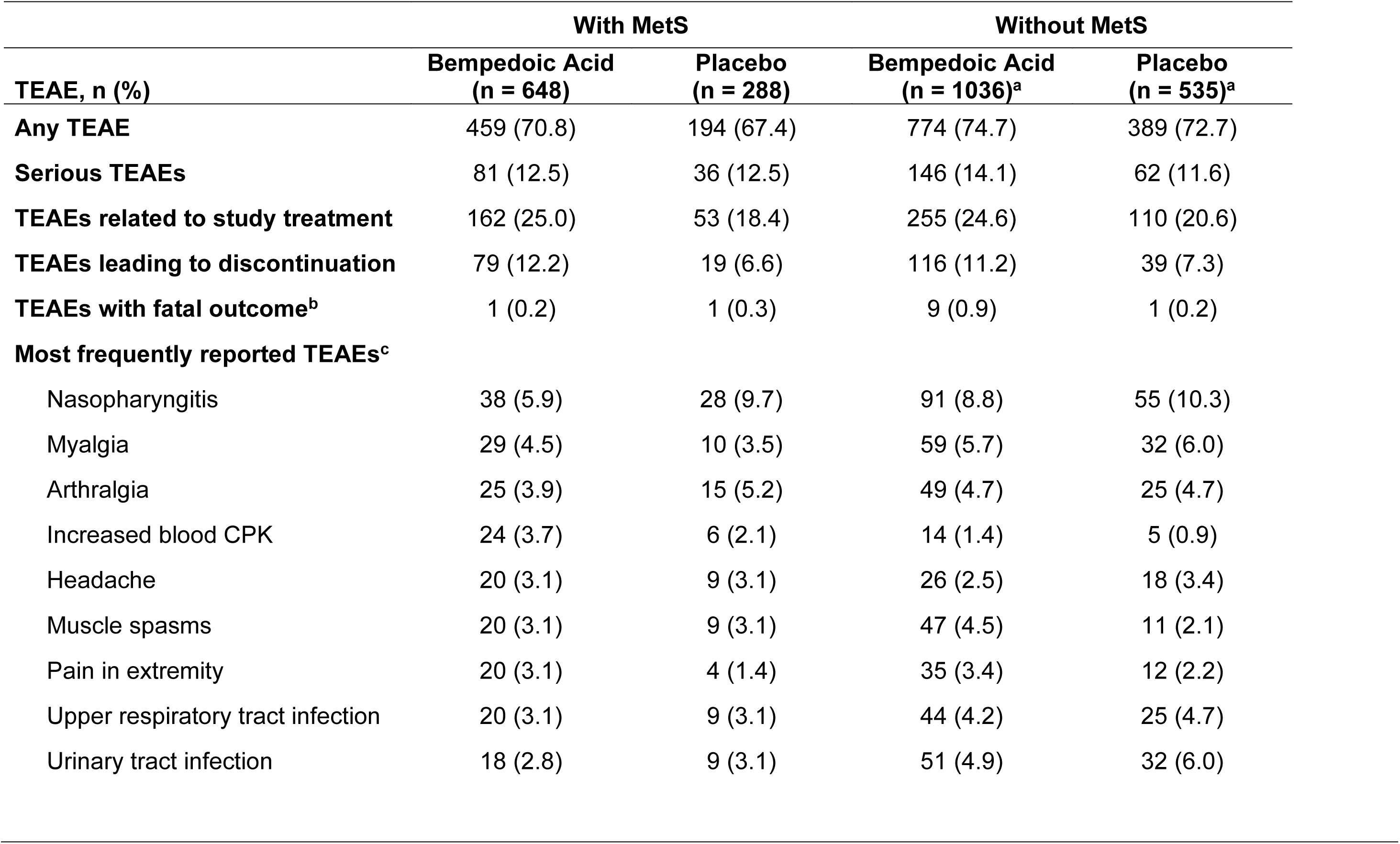
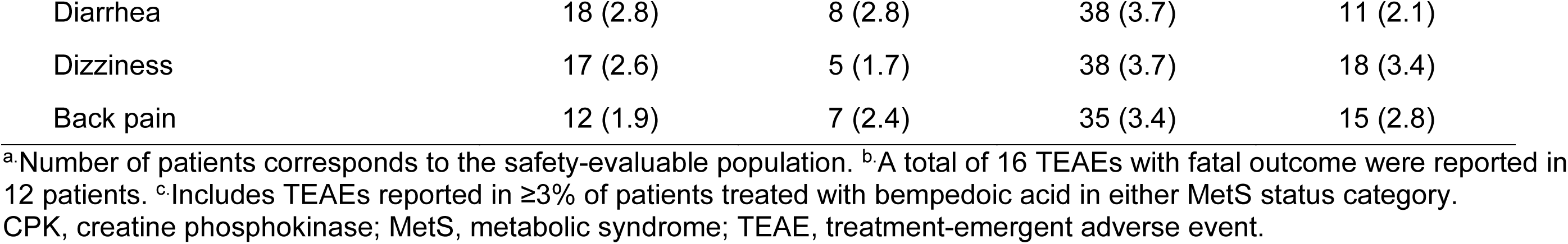
Safety Overview.

|  | With MetS |  | Without MetS |  |
| --- | --- | --- | --- | --- |
| TEAE, n (%) | Bempedoic Acid<br>(n = 648) | Placebo<br>(n = 288) | Bempedoic Acid<br>(n = 1036)<sup>a</sup> | Placebo<br>(n = 535)<sup>a</sup> |
| **Any TEAE** | 459 (70.8) | 194 (67.4) | 774 (74.7) | 389 (72.7) |
| **Serious TEAEs** | 81 (12.5) | 36 (12.5) | 146 (14.1) | 62 (11.6) |
| **TEAEs related to study treatment** | 162 (25.0) | 53 (18.4) | 255 (24.6) | 110 (20.6) |
| **TEAEs leading to discontinuation** | 79 (12.2) | 19 (6.6) | 116 (11.2) | 39 (7.3) |
| **TEAEs with fatal outcome<sup>b</sup>** | 1 (0.2) | 1 (0.3) | 9 (0.9) | 1 (0.2) |
| **Most frequently reported TEAEs<sup>c</sup>** |  |  |  |  |
| Nasopharyngitis | 38 (5.9) | 28 (9.7) | 91 (8.8) | 55 (10.3) |
| Myalgia | 29 (4.5) | 10 (3.5) | 59 (5.7) | 32 (6.0) |
| Arthralgia | 25 (3.9) | 15 (5.2) | 49 (4.7) | 25 (4.7) |
| Increased blood CPK | 24 (3.7) | 6 (2.1) | 14 (1.4) | 5 (0.9) |
| Headache | 20 (3.1) | 9 (3.1) | 26 (2.5) | 18 (3.4) |
| Muscle spasms | 20 (3.1) | 9 (3.1) | 47 (4.5) | 11 (2.1) |
| Pain in extremity | 20 (3.1) | 4 (1.4) | 35 (3.4) | 12 (2.2) |
| Upper respiratory tract infection | 20 (3.1) | 9 (3.1) | 44 (4.2) | 25 (4.7) |
| Urinary tract infection | 18 (2.8) | 9 (3.1) | 51 (4.9) | 32 (6.0) |
| Diarrhea | 18 (2.8) | 8 (2.8) | 38 (3.7) | 11 (2.1) |
| Dizziness | 17 (2.6) | 5 (1.7) | 38 (3.7) | 18 (3.4) |
| Back pain | 12 (1.9) | 7 (2.4) | 35 (3.4) | 15 (2.8) |

<sup>a</sup>Number of patients corresponds to the safety-evaluable population. <sup>b</sup>A total of 16 TEAEs with fatal outcome were reported in 12 patients. <sup>c</sup>Includes TEAEs reported in $\geq 3\%$ of patients treated with bempedoic acid in either MetS status category. CPK, creatine phosphokinase; MetS, metabolic syndrome; TEAE, treatment-emergent adverse event.

Serious TEAEs in patients without MetS were reported in 146 (14.1%) and 62 (11.6%) patients who received bempedoic acid and placebo, respectively. TEAEs related to study treatment were more frequent with bempedoic acid than with placebo regardless of metabolic status, reported in 162 (25.0%) and 53 (18.4%) patients with MetS, respectively, and 255 (24.6%) and 110 (20.6%) patients without MetS, respectively. TEAEs leading to discontinuation of bempedoic acid and placebo were reported in 79 (12.2%) and 19 (6.6%) patients with MetS, respectively, and in 116 (11.2%) and 39 (7.3%) patients without MetS, respectively.

Nasopharyngitis was the most common TEAE in each of the four groups, reported in 38 (5.9%) and 28 (9.7%) patients with MetS and in 91 (8.8%) and 55 (10.3%) patients without MetS, who received bempedoic acid and placebo, respectively (**Table 2**). Among patients treated with bempedoic acid, myalgia was the second-most frequently reported TEAE in each MetS status category, reported in 29 (4.5%) patients with MetS (*vs*. 10 [3.5%] patients who received placebo) and 59 (5.7%), without MetS (*vs*. 32 [6.0%] patients with placebo). Arthralgia was reported in 25 (3.9%) and 15 (5.2%) patients with MetS who received bempedoic acid and placebo, respectively, and in 49 (4.7%) and 25 (4.7%) patients without MetS, respectively. Other TEAEs that were reported at a higher frequency with bempedoic acid than with placebo in patients with MetS included increased blood creatine phosphokinase, reported in 24 (3.7%) patients who received bempedoic acid (*vs*. 6 [2.1%] with placebo) and pain in extremity, reported in 20 (3.1%) patients (*vs*. 4 [1.4%] with placebo). Among patients without MetS, TEAEs reported more frequently with bempedoic acid than placebo included muscle spasms, reported in 47 (4.5%) patients who received bempedoic acid (*vs*. 11 [2.1%] with placebo) and pain in extremity, reported in 35 (3.4%) patients (*vs*. 12 [2.2%] with placebo). Across all groups, a total of 16 TEAEs with fatal outcome were reported in 12 patients (**Table 2** and Supplementary Table 1).

## Discussion

Treatment with bempedoic acid was associated with significant decreases in LDL-C, total cholesterol, non–HDL-C, ApoB, and hsCRP compared with placebo, regardless of metabolic status (*p*<0.0001 for all). While no significant differences in efficacy were observed between patients with and without MetS for total cholesterol, non–HDL-C, ApoB, and hsCRP, patients with MetS treated with bempedoic acid had significantly greater LDL-C reductions than those without MetS (-22.3% *vs*. -18.4%; interaction *p*=0.0472). Among patients treated with bempedoic acid, triglycerides increased from baseline regardless of MetS status. The increase was significantly greater in patients with MetS than in those without, but not to the extent of being clinically meaningful. These results were somewhat unexpected given that bempedoic acid has been shown to have no effect on triglyceride levels *vs*. control in a meta-analysis of eight studies [27], which included two of the studies used for our analyses (CLEAR Harmony [22] and CLEAR Serenity [21]).

Although significant, the reductions with bempedoic acid in LDL-C, non–HDL-C, and ApoB in the current study were smaller than those observed in an analysis of ten phase 3 ODYSSEY trials investigating the efficacy and safety of the PCSK9 inhibitor, alirocumab, in patients with and without MetS [14]. This study, which also excluded patients with diabetes, found significant reductions compared with control in each of these parameters, regardless of MetS status. A significantly greater magnitude of benefit in patients with *vs*. without MetS at week 12 in lowering LDL-C was only observed for pooled data from two studies in which patients were randomized to alirocumab 150 mg or placebo alongside background statins. Significantly greater placebo-corrected reductions in ApoB at week 24, but not at week 12, were also observed in patients with *vs*. without MetS who received alirocumab 75/150 mg. The same study found that triglyceride lowering was only significant for studies in which placebo with background statins was the comparator; no differences were observed between MetS categories.

Significantly greater lowering of lipid parameters beyond LDL-C in patients with *vs*. without MetS has been observed in patients treated with rosuvastatin plus ezetimibe in a phase 3 clinical trial conducted in South Korea [28]. In this study, patients with MetS experienced significantly greater reductions at week 8 in total cholesterol, non– HDL-C, and ApoB, as well as LDL-C, compared with those without MetS. No such differences were observed among patients who received rosuvastatin as monotherapy.

In our study, small statistically significant reductions in HbA1c and FPG with bempedoic acid *vs*. placebo were only observed among patients with MetS, indicative of improved glycemic control with bempedoic acid in these patients compared with those without MetS. In contrast, the PCSK9 inhibitor, alirocumab, has been shown to have no effect on either of these parameters in the pooled analysis of data from ten ODYSSEY trials [14]. Similarly, data from the Further Cardiovascular Outcomes Research With PCSK9 Inhibition in Subjects With Elevated Risk (FOURIER) trial showed that levels of HbA1c at week 48 were similar between patients receiving the PCSK9 inhibitor, evolocumab, and placebo, regardless of metabolic status [15].

Levels of hsCRP, a marker of inflammation, have been shown to be higher in patients with MetS than in those without [29], and high hsCRP levels have been associated with impaired insulin sensitivity [30]. As expected, in our study, patients with MetS had higher baseline hsCRP than those without MetS. The percent decrease in hsCRP from baseline was greater in patients treated with bempedoic acid than in those who received placebo in both MetS status categories, and the magnitude was similar between MetS status categories.

Bempedoic acid was generally well tolerated, with comparable safety profiles in patients with and without MetS. Among patients who received bempedoic acid, the most frequently reported TEAEs in both MetS categories were nasopharyngitis and myalgia, both of which were reported at a similar frequency in patients who received placebo in this study and in the overall pooled patient population [23]. Within each MetS category, the incidence of TEAEs was similar between the bempedoic acid and placebo treatment groups.

A limitation of our study is the retrospective nature of the analyses, which used pooled data from clinical trials that were not originally designed to compare the effects of bempedoic acid in patients with and without MetS. A further limitation is the high proportion of White patients (>95% overall) included in the study, which limits the generalizability of our results. Additional studies evaluating bempedoic acid in patients with or without MetS are therefore needed to validate our findings.

Our results show that bempedoic acid may be used as an adjunct to diet and maximally tolerated statin therapy in patients with and without MetS who require additional lowering of LDL-C and other lipids.

## Data Availability

All data produced in the present study are available upon reasonable request to the authors

## Author Contributions

MJL conceptualized and designed the study. MDS, PRT, and CMB were involved in the data acquisition. LL performed the statistical analysis. All authors participated in data interpretation, critically reviewed and approved the final manuscript draft submitted for publication, and agree to be accountable for all aspects of the work, ensuring the accuracy and integrity of the publication.

## Acknowledgements

The authors would like to thank the patients, their families, and all investigators involved in this study. Medical writing and editorial assistance were provided by David Buist, PhD, and Agata Shodeke, PhD, of Spark Medica Inc., supported by Esperion Therapeutics, Inc., according to Good Publication Practice guidelines. The Sponsor was involved in the study design, collection, and analysis and interpretation of data. However, ultimate responsibility for opinions, conclusions, and data interpretation lies with the authors.

