## Supplementary Material for "Efficacy and Safety of Bempedoic Acid in Patients With and Without Metabolic Syndrome: Pooled Analysis of Data From Four Phase 3 Clinical Trials"

### **Supplementary Table 1. TEAEs With Fatal Outcome**

|  | **With MetS** | | **Without MetS** | |
| --- | --- | --- | --- | --- |
|  | **Bempedoic Acid (n = 648)** | **Placebo (n = 288)** | **Bempedoic Acid (n = 1036)^a^** | **Placebo (n = 535)^a^** |
| **Patients with ≥1 TEAE with fatal outcome,^b^ n (%)** | 1 (0.2) | 1 (0.3) | 9 (0.9) | 1 (0.2) |
| **TEAEs with fatal outcome, n (%)** |  |  |  |  |
| Hypertensive heart disease | 1 (0.2)^c^ | 0 | 0 | 0 |
| Myocardial ischemia | 1 (0.2)^c^ | 0 | 0 | 0 |
| Peritonitis | 0 | 1 (0.3)^d^ | 0 | 0 |
| Septic shock | 0 | 1 (0.3)^d^ | 0 | 0 |
| Chronic obstructive pulmonary disease | 0 | 0 | 1 (0.1)^e^ | 0 |
| Malignant lung neoplasm | 0 | 0 | 1 (0.1)^e^ | 0 |
| Multiple organ dysfunction syndrome | 0 | 0 | 1 (0.1)^f^ | 0 |
| Sepsis | 0 | 0 | 1 (0.1)^f^ | 0 |
| Pancreatic pseudocyst | 0 | 0 | 1 (0.1) | 0 |
| Ischemic cerebral infarction | 0 | 0 | 1 (0.1) | 0 |
| Lung adenocarcinoma | 0 | 0 | 1 (0.1) | 0 |
| Cardiac arrest | 0 | 0 | 1 (0.1) | 0 |
| Gas poisoning | 0 | 0 | 1 (0.1) | 0 |
| Myocardial infarction | 0 | 0 | 1 (0.1) | 0 |
| Death | 0 | 0 | 1 (0.1) | 0 |
| Acute coronary syndrome | 0 | 0 | 0 | 1 (0.2) |

^a.^Number of patients corresponds to the safety-evaluable population. ^b.^A total of 16 TEAEs with fatal outcome were reported in 12 patients. ^c.^Hypertensive heart disease and myocardial ischemia with fatal outcome were both reported in the same patient. ^d.^Peritonitis and septic shock with fatal outcome were both reported in the same patient. ^e.^Chronic obstructive pulmonary disease and malignant lung neoplasm with fatal outcome were both reported in the same patient. ^f.^Multiple organ dysfunction syndrome and sepsis were both reported in the same patient.
MetS, metabolic syndrome; TEAE, treatment-emergent adverse event.
